# Non-existent ClinicalTrials.gov identifiers in abstracts indexed by PubMed

**DOI:** 10.1101/2020.02.24.20027300

**Authors:** Benjamin Gregory Carlisle

## Abstract

Prospective registration plays an important role in ensuring the transparency and reliability of clinical trials. Preregistration of clinical trials has been required by the ICMJE since 2005 and mandated by law for most clinical trial types since 2007. It is one of the roles of peer reviewers of a clinical trial publication to confirm that there is concordance between the registry entry and the submitted publication. On October 22, 2019, abstracts for all articles indexed by PubMed with publication type “Clinical Trial” and a publication date after January 1, 2003 were downloaded. Clinical trial registry identifiers were automatically extracted and tested for the existence of a corresponding entry on ClinicalTrials.gov. Among 38,001 published clinical trial registry numbers, 215 (0.6%) do not correspond to a legitimate clinical trial registry entry. While there is a small proportion of non-existent NCT numbers in our sample, even a single non-existent NCT number in a publication represents a failure on the part of journals who publish clinical trials to systematically ensure that reviewers always check clinical trial registry entries for concordance with the text submitted for publication. These results cast doubt on how frequently editors and reviewers evaluate clinical trial reports in light of their corresponding registry entries.

## Background

Preregistration of clinical trials plays an important part in preventing selective reporting of outcomes, switching of primary and secondary endpoints, and certain kinds of publication bias.^1,2^ Prospective registration also allows for greater transparency in the conduct and reporting of clinical trials,^3^ which can bolster confidence in the proper functioning of the machinery of human research. Prospective clinical trial registration has been required by the ICMJE since 2005^4^ and has been legally mandated in the United States for most clinical trial types since 2007.^5^

## Methods

On October 22, 2019, all articles indexed by PubMed with publication type “Clinical Trial” and a publication date after January 1, 2003 were downloaded. The PubMed ID, date of publication, abstract and journal name were extracted from the PubMed XML metadata. Reported trial registration (“NCT”) numbers were automatically extracted from abstracts and checked for the existence of a corresponding record on ClinicalTrials.gov. NCT numbers that did not exist on ClinicalTrials.gov were verified manually to ensure that they had been extracted correctly from the abstract text and that the registry entry does not exist. The complete data set and code for extraction of NCT numbers and checking for a corresponding record on ClinicalTrials.gov are available online.^6,7^

## Results

Among 488,364 clinical trial publications indexed by PubMed, there were 38,001 published NCT numbers. Of these NCT numbers, 215 (0.6%) do not correspond to a clinical trial registry entry on ClinicalTrials.gov; see Figure 1.

**Figure 1.**
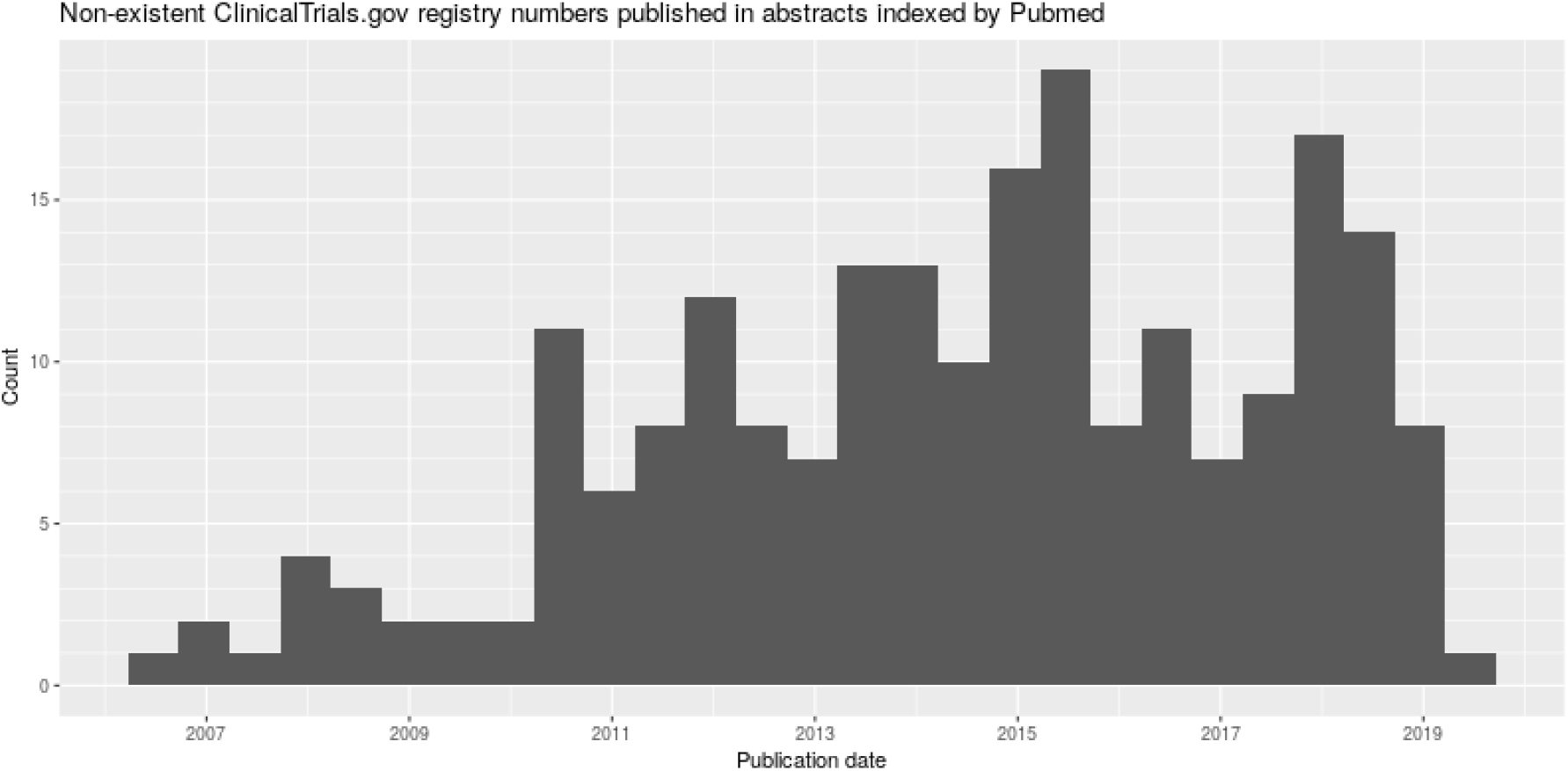
Non-existent ClinicalTrials.gov registry numbers published in abstracts indexed by PubMed

The abstracts containing non-existent NCT numbers were published by 132 distinct journals which published an average of 577 clinical trials in our sample (range 7-7315). These journals reported an average of 134 NCT numbers in our sample (range 1-1696). For 9 of the 215 non-existent NCT numbers (4%), there was a separate, secondary publication of that clinical trial in our sample that also referred to the same non-existent NCT number.

In a random sample of 22 (10%) of the 215 extracted NCT numbers, a manual search based on trial details (drug, indication, etc.) or other identifiers (e.g. non-NCT trial identifier) in the abstract or the full-text of the publication found that all 22 do have a valid corresponding record, either in ClinicalTrials.gov or another public registry. Seventeen (77%) of the extracted NCT numbers differed from the actual NCT number by only a single digit. In two cases, an identifier for another public registry was incorrectly reported as a ClinicalTrials.gov NCT number. Among the random sample of 22 NCT numbers, 8 of the non-existent NCT numbers were repeated in the full text, 10 had no NCT number reported in the full text at all, and there were 4 cases where the non-existent NCT number appeared only in the abstract and a legitimate NCT number was presented in the full article text. See data set for details.^7^

## Discussion

While there is a small proportion of non-existent NCT numbers in clinical trial abstracts indexed by PubMed, publication of even a single non-existent NCT number represents a failure on the part of journals who publish clinical trials to systematically ensure that clinical trial registry entries are always checked for concordance with the trial report submitted for publication.

Because we were able to identify legitimate registry entries for the entire random sample that we checked manually, and in a small number of cases, the correct NCT number was reported in the full text publication, these errors appear to be simple data entry mistakes on the part of the authors, rather than a malicious attempt to hide a clinical trial registry from scrutiny. The journals implicated here are not predatory journals, but rather reputable, high-impact journals such as *The Lancet*,^8–11^*Blood*,^12–18^*PLOS One*,^19–23^*BMJ*^24,25^*Circulation*,^26–29^ and *Trials*.^30–34^

In the 215 cases of non-existent NCT numbers that were identified, because the error was not corrected during the publication process, and because our results suggest that there are few cases in which the full publication text contains an accurate registry number when the number in the abstract does not exist, it is very likely that none of the editors or reviewers attempted to even access the clinical trial registry entry. If this is the case, peer review and editorial scrutiny provided no evaluation at all of the concordance between the details provided on the ClinicalTrials.gov record, such as primary and secondary outcomes, and those reported in the publication. This is consistent with what others have reported regarding peer review,^35^and suggests that editors and others involved in journal publication also do not review every clinical trial for concordance with the reported trial registry entry. The finding of even a single non-existent NCT number in a publication, to say nothing of serial publications of non-existent NCT numbers referring to the same clinical trial, suggests that there is no mechanism ensuring confirmation of clinical trial registry details by editors and reviewers of clinical trial publications, and casts doubt on how frequently editors and reviewers evaluate clinical trial reports in light of their corresponding registry entries.

A survey of 203 peer-reviewers found that verification of trial registration details was never clearly requested by journal editors.^36^The problem of published NCT numbers that do not correspond to a legitimate ClinicalTrials.gov record could be resolved by the introduction of a check-list to be used by reviewers and editors requiring them to confirm concordance of important clinical trial registry details with the publication text. It would even be possible to introduce an automated tool for reviewers that identifies numbers within a manuscript that are formatted as a clinical trial registry entry and retrieves details of a clinical trial registry entry from the text of a paper or abstract, or in the case of non-existent ones, flags them for correction.

This study is limited in that for feasibility, this sample only includes registry numbers extracted from abstracts indexed by PubMed. Clinical trials may also report non-existent NCT numbers in the full text of the article.^37^Hence, this analysis places a lower bound on the number of published NCT numbers that do not correspond to a legitimate ClinicalTrials.gov registry entry.

One of the main ways that clinical trial preregistration allows for accountability is at the point of trial publication. Even in cases where a clinical trial is pre-registered, inconsistencies such as outcome-switching between the registered trial and the publication are possible. It is the responsibility of peer reviewers of a clinical trial publication to confirm that there is concordance between the clinical trial registry entry data and the submitted text for publication. The role of clinical trial registration in holding investigators accountable is compromised if journal editors and peer reviewers do not refer to clinical trial registries in order to ensure that there is concordance between the registry record and the publication text.

END

## Data Availability

The data referred to are available from the Open Science Foundation

https://dx.doi.org/10.17605/OSF.IO/HFM9Y

## Acknowledgements

Many thanks to Alex Bannach-Brown, Peter Grabitz and Jonathan Kimmelman for conversations that inspired this paper; and to Maia Salholz-Hillel for insightful feedback.

## References

1. Simes, R. J. Publication bias: The case for an international registry of clinical trials. Journal of clinical oncology 4, 1529–1541 (1986).

2. To, M. J., Jones, J., Emara, M. & Jadad, A. R. Are reports of randomized controlled trials improving over time? A systematic review of 284 articles published in high-impact general and specialized medical journals. PLoS One 8, (2013).

3. Nosek, B. A., Ebersole, C. R., DeHaven, A. C. & Mellor, D. T. The preregistration revolution. Proceedings of the National Academy of Sciences 115, 2600–2606 (2018).

4. De Angelis Catherine et al. Clinical Trial Registration. Circulation 111, 1337–1338 (2005).

5. Food and Drug Administration Amendments Act of 2007.

6. Carlisle, B. G. Pubmed-NCT-extractor. (2020). Available at: https://codeberg.org/bgcarlisle/Pubmed-NCT-extractor. (Accessed: 4th February 2020)

7. Carlisle, B. G. Non-existent ClinicalTrials.Gov identifiers in abstracts indexed by PubMed. (2020). doi:10.17605/OSF.IO/HFM9Y

8. Yousafzai, A. K., Rasheed, M. A., Rizvi, A., Armstrong, R. & Bhutta, Z. A. Effect of integrated responsive stimulation and nutrition interventions in the Lady Health Worker programme in Pakistan on child development, growth, and health outcomes: A cluster-randomised factorial effectiveness trial. Lancet 384, 1282–1293 (2014).

9. Zinman, B. et al. Low-dose combination therapy with rosiglitazone and metformin to prevent type 2 diabetes mellitus (CANOE trial): A double-blind randomised controlled study. Lancet 376, 103–111 (2010).

10. Wenzel, S., Wilbraham, D., Fuller, R., Getz, E. B. & Longphre, M. Effect of an interleukin-4 variant on late phase asthmatic response to allergen challenge in asthmatic patients: Results of two phase 2a studies. Lancet 370, 1422–1431 (2007).

11. Catovsky, D. et al. Assessment of fludarabine plus cyclophosphamide for patients with chronic lymphocytic leukaemia (the LRF CLL4 Trial): A randomised controlled trial. Lancet 370, 230–239 (2007).

12. Santagostino, E. et al. Long-acting recombinant coagulation factor IX albumin fusion protein (rIX-FP) in hemophilia B: Results of a phase 3 trial. Blood 127, 1761–1769 (2016).

13. Sun, C. et al. Partial reconstitution of humoral immunity and fewer infections in patients with chronic lymphocytic leukemia treated with ibrutinib. Blood 126, 2213–2219 (2015).

14. Rook, A. H. et al. Topical resiquimod can induce disease regression and enhance T-cell effector functions in cutaneous T-cell lymphoma. Blood 126, 1452–1461 (2015).

15. Johnson, G. G. et al. CYP2B6*6 is an independent determinant of inferior response to fludarabine plus cyclophosphamide in chronic lymphocytic leukemia. Blood 122, 4253–4258 (2013).

16. Scully, M. et al. A phase 2 study of the safety and efficacy of rituximab with plasma exchange in acute acquired thrombotic thrombocytopenic purpura. Blood 118, 1746–1753 (2011).

17. Burt, R. K. et al. Autologous nonmyeloablative hematopoietic stem cell transplantation in patients with severe anti-TNF refractory Crohn disease: Long-term follow-up. Blood 116, 6123–6132 (2010).

18. Cassani, B. et al. Altered intracellular and extracellular signaling leads to impaired T-cell functions in ADA-SCID patients. Blood 111, 4209–4219 (2008).

19. Ayieko, J. et al. “Hurdles on the path to 90-90-90 and beyond”: Qualitative analysis of barriers to engagement in HIV care among individuals in rural East Africa in the context of test-and-treat. PLoS ONE 13, e0202990 (2018).

20. Somé, E. N. et al. Changes in body mass index and hemoglobin concentration in breastfeeding women living with HIV with a CD4 count over 350: Results from 4 African countries (The ANRS 12174 trial). PLoS ONE 12, e0177259 (2017).

21. Andersson, E. et al. Genetic polymorphisms in monoamine systems and outcome of cognitive behavior therapy for social anxiety disorder. PLoS ONE 8, e79015 (2013).

22. Cohen, C. R. et al. A phase I randomized placebo controlled trial of the safety of 3% SPL7013 Gel (VivaGel®) in healthy young women administered twice daily for 14 days. PLoS ONE 6, e16258 (2011).

23. Peters, R. et al. Modelling cognitive decline in the Hypertension in the Very Elderly Trial [HYVET] and proposed risk tables for population use. PLoS ONE 5, e11775 (2010).

24. Myers, M. G. et al. Conventional versus automated measurement of blood pressure in primary care patients with systolic hypertension: Randomised parallel design controlled trial. BMJ 342, d286 (2011).

25. Picado, A. et al. Longlasting insecticidal nets for prevention of Leishmania donovani infection in India and Nepal: Paired cluster randomised trial. BMJ 341, c6760 (2010).

26. Barst, R. J. et al. STARTS-2: Long-term survival with oral sildenafil monotherapy in treatment-naive pediatric pulmonary arterial hypertension. Circulation 129, 1914–1923 (2014).

27. Roe, M. T. et al. Elderly patients with acute coronary syndromes managed without revascularization: Insights into the safety of long-term dual antiplatelet therapy with reduced-dose prasugrel versus standard-dose clopidogrel. Circulation 128, 823–833 (2013).

28. Herrmann, H. C. et al. Predictors of mortality and outcomes of therapy in low-flow severe aortic stenosis: A Placement of Aortic Transcatheter Valves (PARTNER) trial analysis. Circulation 127, 2316–2326 (2013).

29. Rodés-Cabau, J. et al. Comparison of plaque sealing with paclitaxel-eluting stents versus medical therapy for the treatment of moderate nonsignificant saphenous vein graft lesions: The moderate vein graft lesion stenting with the taxus stent and intravascular ultrasound (VELETI) pilot trial. Circulation 120, 1978–1986 (2009).

30. Batista, K. B. D. S. L. et al. Herbst appliance with skeletal anchorage versus dental anchorage in adolescents with Class II malocclusion: Study protocol for a randomised controlled trial. Trials 18, 564 (2017).

31. Wickwar, S. et al. Effectiveness and cost-effectiveness of a patient-initiated botulinum toxin treatment model for blepharospasm and hemifacial spasm compared to standard care: Study protocol for a randomised controlled trial. Trials 17, 129 (2016).

32. Nyende, S. et al. Solar-powered oxygen delivery: Study protocol for a randomized controlled trial. Trials 16, 297 (2015).

33. Calamita, S. A. P. et al. Evaluation of the immediate effect of acupuncture on pain, cervical range of motion and electromyographic activity of the upper trapezius muscle in patients with nonspecific neck pain: Study protocol for a randomized controlled trial. Trials 16, 100 (2015).

34. Huang, K.-Y. et al. Implementing an early childhood school-based mental health promotion intervention in low-resource Ugandan schools: Study protocol for a cluster randomized controlled trial. Trials 15, 471 (2014).

35. Mathieu, S., Chan, A.-W. & Ravaud, P. Use of trial register information during the peer review process. PLoS One 8, (2013).

36. Chauvin, A., Ravaud, P., Baron, G., Barnes, C. & Boutron, I. The most important tasks for peer reviewers evaluating a randomized controlled trial are not congruent with the tasks most often requested by journal editors. BMC Med 13, 158 (2015).

37. Renouf, D. J. et al. A phase II study of the halichondrin B analog eribulin mesylate in gemcitabine refractory advanced pancreatic cancer. Investigational New Drugs 30, 1203–1207 (2012).

